# Are pregnant women satisfied with perinatal standards of care during COVID-19 pandemic?

**DOI:** 10.1101/2020.11.19.20231670

**Authors:** Claudia Ravaldi, Laura Mosconi, Giada Crescioli, Valdo Ricca, Alfredo Vannacci

**Affiliations:** CiaoLapo Foundation for Perinatal Health, Prato, Italy; PeaRL Perinatal Research Laboratory, University of Florence; Department of Neurosciences, Psychology, Drug Research and Child Health, University of Florence, Florence, Italy; Department of Health Sciences, University of Florence, Florence, Italy

## Abstract

COVID-19 restrictive measures severely impacted maternity services worldwide, but little is known about the differences in women’s concerns, perception of the modifications of maternity services and childbirth programs at different times during the pandemic. Here we report data from the first COVID-19 wave in Italy, during the 2020 national lockdown (March-April) and soon after lockdown release (May).

1307 pregnant women answered the survey during national lockdown (phase 1) or after restrictive measures were released (phase 2). Women reported a significantly higher COVID-19 concern during phase 1 than during phase 2 (2.34 SD 0.5 vs 2.12 SD 0.5 on a Likert scale 0-3; p<0.001). Several domains of perinatal care were affected during COVID-19 lockdown: while antenatal visits, the use of technology to keep in touch with healthcare professionals, and closeness of caregivers were generally more appreciated (especially during phase 2), women reported the greatest difficulties in receiving clear information on hospitalization, birth plan and partner’s presence at birth.

Italian pregnant women’s worries about the effects of the pandemic on health and their perception of quality in the organization of maternity services improved during lockdown, but they continued to represent a challenge in May, especially regarding organizational aspects of hospitalization and childbirth.

## Introduction

Since January 30^th^ 2020, the World Health Organization (WHO) declared the pandemic status due to COVID-19. In order to contain the spread of the infection, restrictive measures have been adopted in many countries and in particular in Italy, the first COVID-19 pandemic epicenter among European countries. Italian government established a period of full ‘lockdown’, consisting of travel bans, mandatory staying at home and temporary closure of non-essential businesses. Furthermore, first weeks of the epidemic were also characterised by a shortage of personal protective equipment [1], so that access to health services needed to be significantly reorganised, with a limitation of visits in person and postponement of all non-urgent appointments. The reorganisation of services severely impacted maternity services worldwide [2], so that at the beginning of March, WHO highlighted the need of maintaining respectful care during pregnancy, labour and childbirth despite the pandemic for all women, including those who were affected by COVID-19 or tested positive to SARSCoV2 infection; this position was enforced internationally both by professionals and by human rights advocates [3, 4]. Studies conducted during the first months of the pandemic highlighted that pregnant women suffered for health-related restrictions, were worried about the lack of clear and detailed information from their maternity services and expressed concern about their labour and birth [5]. After a few months of strict restrictions, virus spread decreased almost everywhere and many activities and services resumed. In Italy, lockdown was released at the beginning of May and, despite the maintenance of an “emergency status”, maternity services were allowed to reorganise their activities without the restrictions they had adopted during the pandemic. We have already shown that during the very first weeks of COVID-19 pandemic childbirth scenario was very frightening for Italian women [5], and this condition was also shown to negatively influence their mental health [6]. Nevertheless, little is known on what actually changed after lockdown measures were released.

In order to address the differences in women’s perception of maternity services organization and childbirth programs that were adopted at different times during the pandemic, here we report data collected in the first COVID-19 wave in Italy during the 2020 national lockdown (March-April) and soon after lockdown release (after May 3^rd^).

## Methods

Detailed methods of the study have already been described [7]. Briefly, the nation-wide cross-sectional web-based survey COVID-ASSESS was conducted between March and May 2020 in a sample of Italian pregnant women, recruited with a snowball technique.

The questionnaire recorded sociodemographic and anamnestic variables, women’s childbirth expectations, their concerns about COVID-19, and their perception of information and communication by healthcare providers during the epidemic. Human research ethical approval to conduct the survey was received from Florence University ethics committee (prot. n. 006897).

## Results

A total of 1307 pregnant women answered the survey during national lockdown (phase 1) or after restrictive measures were released (phase 2). Throughout Italy, women reported a significantly higher concern for COVID-19 during phase 1 than during phase 2 (overall mean concern 2.34, SD 0.5 vs 2.12, SD 0.5 on a Likert scale 0-3; p<0.001; Fig. 1A).

**Figure 1.**
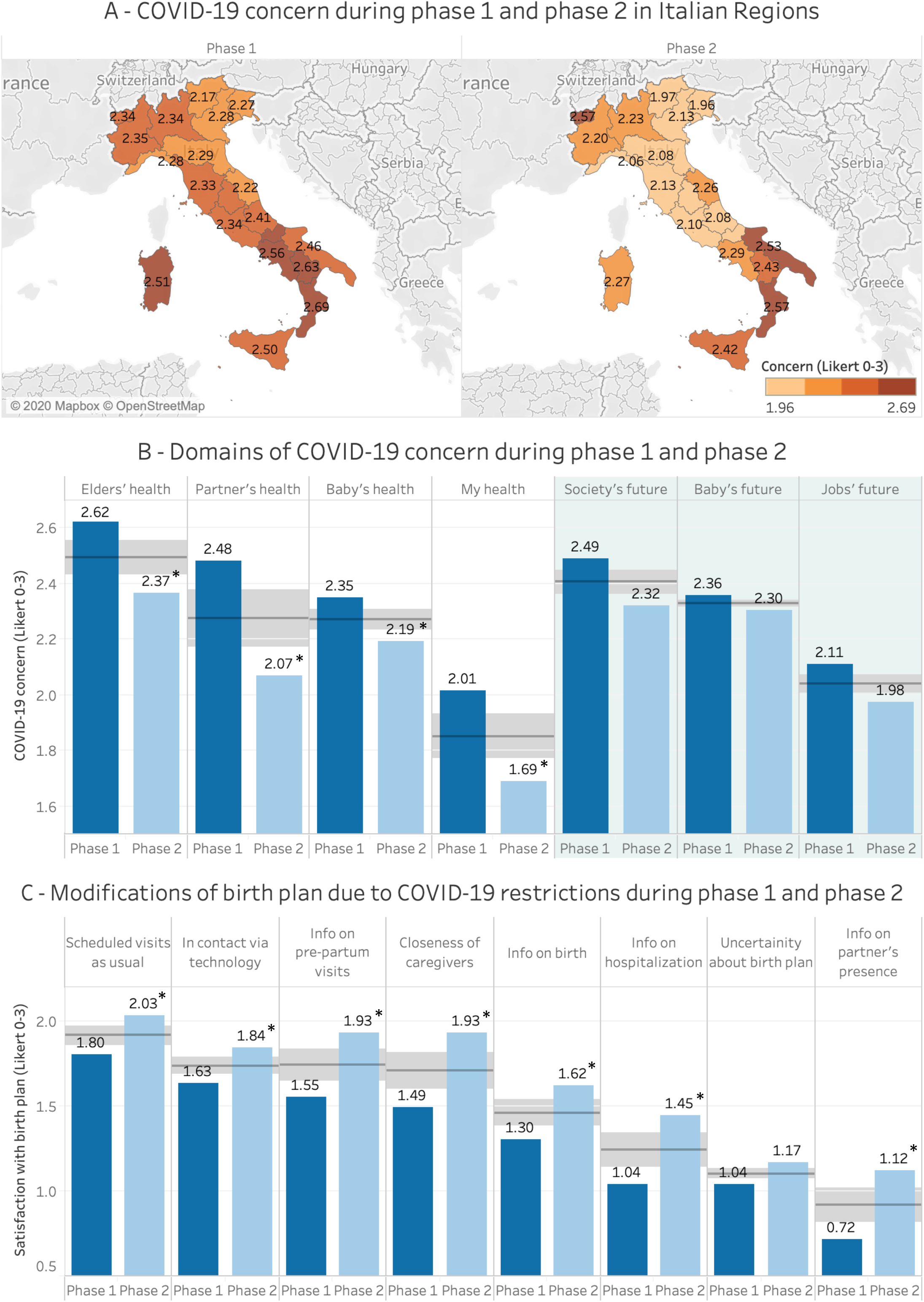
Distribution of COVID-19 concern in Italian regions during phase 1 and phase 2 (panel A). Domains of COVID-19 concern in health issues (white background) and future issues (grey background) during phase 1 and phase 2 (panel B). Satisfaction of pregnant women with the modifications of perinatal standards of care during phase 1 and phase 2 (panel C). Horizontal lines in panels B and C represent median and shaded areas represent quartiles. * p<0.01 vs phase 1.

In keeping with a preliminary analysis [5] conducted on 200 subjects recruited during phase 1, pregnant women were significantly less concerned with their own health than with the one of their relatives; concerns with the future (in particular for their baby, their job or society in general) were also high (Fig 1B). In addition, while health concerns significantly decreased during phase 2, concerns for the future did not change (Fig 1B). As expected, several domains of perinatal care were affected during COVID-19 lockdown: while antenatal visits, the use of technology to keep in touch with healthcare professionals, and closeness of caregivers were generally more appreciated (especially during phase 2), women reported the greatest difficulties in receiving clear information on hospitalization, birth plan and partner’s presence at birth (Fig 1C). These difficulties were high during lockdown and improved significantly during the following months, but the majority of women in May were still reporting uncertainty and the lack of a clear organization in several domains of maternity services.

## Discussion

Italian pregnant women’s worries about the effects of the pandemic on health significantly decreased from March 2020 to May 2020. On the one hand, this could be due to a reorganisation of maternity services after the first months of the emergency, on the other hand it could represent an adaptive response of women to stress with the adoption of coping mechanisms after the acute phase of the epidemic. Consistently, the quality perception of the organisation of maternity services also improved, with women reporting higher satisfaction than in phase 1 regarding visits’ information and scheduling, use of technology by their attending healthcare professionals and closeness of caregivers. Despite these improvements, in May organisation of maternity services continued to represent a challenge, especially regarding operational aspects of hospitalization and childbirth. In particular, during phase 2 women still received poor information on their hospitalization programs and on the possibility of fulfilling their birth plan as it was expected before pandemic.

In particular, women’s perception of proper information on their partner presence during labour and childbirth represented the main issue of uncertainty during and soon after COVID-19 first wave in Italy, although the satisfaction increased from March to May. Despite the well-recognised importance of respectful care in such a challenging time [4, 8], the presence of partner or companion of choice during current pandemic at antenatal visits, labour and childbirth still remains controversial in many countries. Activists for human rights in childbirth started a call for action to put respect in pregnancy and childbirth at the centre of care, notwithstanding COVID-19; for example, with the #butnotmaternity campaign women are trying to highlight disparities on pandemic-related restrictions and asking that everybody is allowed a support figure with them for maternal services. Also the Royal College of Midwives and the Royal College of Obstetricians and Gynaecologists recently published a statement on the need of reintroducing partners into maternity care, and the NHS released a promising document soon after that [9]. In Italy as well, the National Institute of Health (Istituto Superiore di Sanità) published a statement to remind all Italian maternity services the need of combining respectful care with the strategies for containing pandemic [10].

In conclusion, despite many global and local efforts, respectful care in maternity service during COVID-19 pandemic seems still to be fragmentary: partners are perceived as visitors and not as caregivers, and not all maternal services have been reorganised. Indeed, this was the situation in Italy during lockdown and soon after the first wave of COVID-19 ended. Further studies are currently ongoing to assess whether or not these issues are being addressed during the second wave of the pandemic and to evaluate the impact of worries related to uncertainty in hospitalization programs and birth plan respect on women’s wellbeing and mental health during pregnancy and postnatal period.

## Data Availability

Original data are available and open.

https://data.mendeley.com/datasets/cn38pbwn7r/1

## Notes

### Competing Interest Statement

The authors have declared no competing interest.

### Clinical Protocols

https://www.sciencedirect.com/science/article/pii/S2352340920313226

### Funding Statement

Acknowledgments
The study was not funded; no researcher received grants, salary or reimbursements for the realization of the study. CiaoLapo Foundation for Healthy Pregnancy and Perinatal Loss Support provided infrastructure for the realization of the study (documents, questionnaires, material, software, web platforms, open access etc).

### Author Declarations

Human research ethical approval to conduct the survey was received from Florence University ethics committee (Prot. n. 006897). Each participant gave their explicit consent in an online form before enrolment.

